# Volatile Organic Compounds for the Detection of Hepatocellular Carcinoma – a Systematic Review

**DOI:** 10.1101/2022.11.14.22282312

**Authors:** Sayed Metwaly, Alicja Psica, Opeyemi Sogaolu, Irfan Ahmed, Ashis Mukhopadhya, Mirela Delibegović, Mohamed Bekheit

**Affiliations:** Division of Internal Medicine, Aberdeen Royal Infirmary, NHS Scotland, United Kingdom; School of Medicine, Medical Sciences and Nutrition, University of Aberdeen, United Kingdom; Department of Critical Care Medicine, University of Calgary, Alberta, Canada; Department of General Surgery, Aberdeen Royal Infirmary, NHS Scotland, United Kingdom; Department of Surgical Oncology. Shaukat Khanum Memorial Cancer Hospital and Research Centre. Lahore, Pakistan; Hépatica, Centre of Integrated HPB Care, Elite Hospital, Alexandria, Egypt

**Author notes:** **Address correspondence to:** Mohamed Bekheit MBChB, MSc, MS Chir, Dip-Biostat, DU-US, FRCS, PhD, Department of General Surgery, NHS Grampian, Foresterhill Health Campus, Foresterhill Rd, Aberdeen AB25 2ZN. **Authors’ contributions:** SM and AP contributed equally to the manuscript and are both the first authors. AP drafted the initial introduction, methods and results section. SM wrote the results, discussion, and introduction and critically reviewed the manuscript. MB conceptualised the study and contributed, with AM, MD, IA, OS to the critical review of the final manuscript. All authors read and approved the final manuscript.

**Keywords:** Hepatocellular carcinoma (HCC), volatile organic compounds (VOCs), metabolomics

## Abstract

**Background:** Hepatocellular carcinoma (HCC) is an increasingly common and one of the leading causes of cancer mortality worldwide. Only a small percentage of HCC patients are eligible to curative treatment. There is a need for a point of care, early diagnostic or screening tool. It is not clear whether exhaled volatile organic compounds (VOCs) could fulfil those needs.

**Hypothesis:** We postulate that exhaled VOCs can identify potential biomarkers for non-invasive detection of HCC.

**Aims:** This systematic review aims to critically review the current knowledge regarding the exhaled VOCs linked to HCC detection.

**Methods:** A systematic electronic search was conducted. Search strategy included all studied published until the 24th of March 2021 using a combination of relevant keywords.

**Results:** The search yielded 6 publications using the PRISMA pathway. Two of the studies described in vitro experiments, and four clinical studies were conducted on small groups of patients. Overall, 42 headspace gases were analysed in the in vitro studies. Combined, the clinical studies included 164 HCC patients and 260 controls. The studies reported potential role for a combination of VOCs in the diagnosis of HCC. However, only limonene, acetaldehyde and ethanol could be traced back to their biological pathways using KEGG pathway enrichment analysis.

**Conclusions:** Although there appears to be promise in VOCs research associated with HCC, there is no single volatile biomarker in exhaled breath attributed to HCC and data from extracted studies indicates a lack of standardization. Large population studies are required to verify the existence of VOCs linked to HCC.

## Introduction

Liver cancer accounted for the fifth cause of cancer-related mortality in males, with 30,200 new cases in the United States in 2019 (1). According to the Global Cancer Statistics 2020, worldwide primary liver cancer has been diagnosed in over 920,000 people and represented third cancer-related death in 2020 (2). The incidence is similar to the mortality rate with 9.5 and 8.7 cases per 100 000, respectively (1). The vast majority of primary liver cancers, nearing 80%, is hepatocellular carcinoma (HCC), and the global incidence has tripled during the past 40 years (3). HCC most commonly arises in the background of liver cirrhosis, and 80 to 90% of patients are found to have cirrhotic changes at the time of diagnosis (4). The aetiology of cirrhosis varies depending on geographical location, however, most prevalent risk factors are HBV and HCV infection, excessive chronic consumption of alcohol, and diabetes or obesity-related NASH (5). Up to one-third of patients with established cirrhosis will be diagnosed with HCC during their lifetime, with an annual incidence rate of 1 to 8% (6). It is crucial to detect liver parenchyma growths early as patients with stage 0, according to Barcelona Clinic Classification, have 70-90% chance of 5-year survival. However, life expectancy dramatically drops to only 4-6 months with the progression of the disease to stage 4 (5).

HCC develops insidiously, and frequently it is diagnosed late. Given the dismal prognosis in advanced cases, several surveillance programs are recommended by national and international bodies such as AASLD, EASL and NICE in the UK (7–9). Those guidelines universally support a regime of 6 monthly USS. Combination with serum Alpha-Fetoprotein (AFP) increases the sensitivity of detection in the early stages of hepatic lesions (10).

AFP is the most widely used HCC biomarker, with a sensitivity of 47.7% and specificity of 97.1% at a cut-off value of 200ng/ml (11).

Considering the rising HCC incidence, research efforts focus on developing new non-invasive, cost and time-effective, population-based screening tests. The association between diseases and changes in breath VOCs has been a matter of heightened interest since antiquity. This is primarily evidenced in liver disease by the clinical observation of foetor hepaticus -a characteristic breath odour (18,19). Recently, numerous pieces of literature published on successfully profiling VOCs in malignancies, such as lung (20,21), breast and colorectal (22), suggest there is a scope for building similar diagnostic tools in liver disease. This approach has been explored in liver cirrhosis (23), however, data about volatilome specific for HCC is generally scarce (12–17). In this systematic review, we aim to compile and critically review existing knowledge related to the utility of VOCs in diagnosing HCC..

## Methodology

This review was conducted according to the Preferred Reporting Items for Systematic Reviews and Meta-analyses (PRISMA) guidelines.

### Search strategy

Two independent authors searched electronic databases PubMed, Medline, OVID Embase and Web of science for articles published before 24^th^ of March 2021. The same search has been performed by an experienced medical librarian from the Royal College of Surgeon of Edinburgh library. Combination of the following key words were used for searching HCC, hepatocellular carcinoma, volatile organic compounds. Keywords were searched in “all fields” and MeSH Term sections. No restriction on language was imposed. Animal studies were excluded as the review focused on human HCC markers. All pertinent publications, excluding conference papers, were analysed.

Additionally, bibliographies of identified studies were searched for relevant publications. Subsequently, candidate articles were scrutinised by two independent investigators according to the PRISMA protocol. A total of 4075 records were identified (Supplementary material – Table S1). Following exclusion of duplicates, rejection of irrelevant papers after review of titles and abstracts and implementation of exclusion criterion of animal studies and conference abstracts, six publications remained for analysis (enumerated in Table 1). Studies selection flow diagram is illustrated in [Fig 1].

**Table 1.**
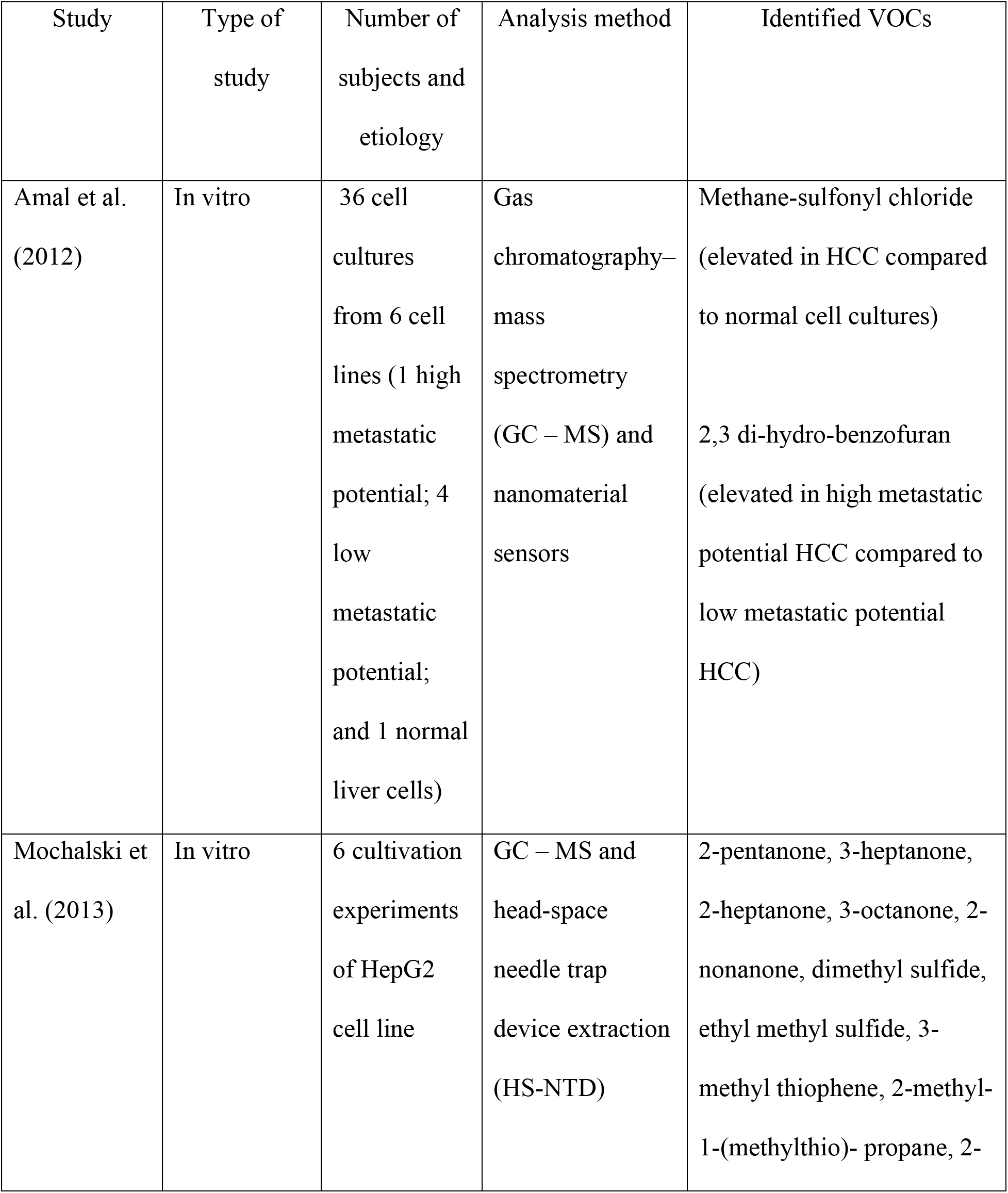
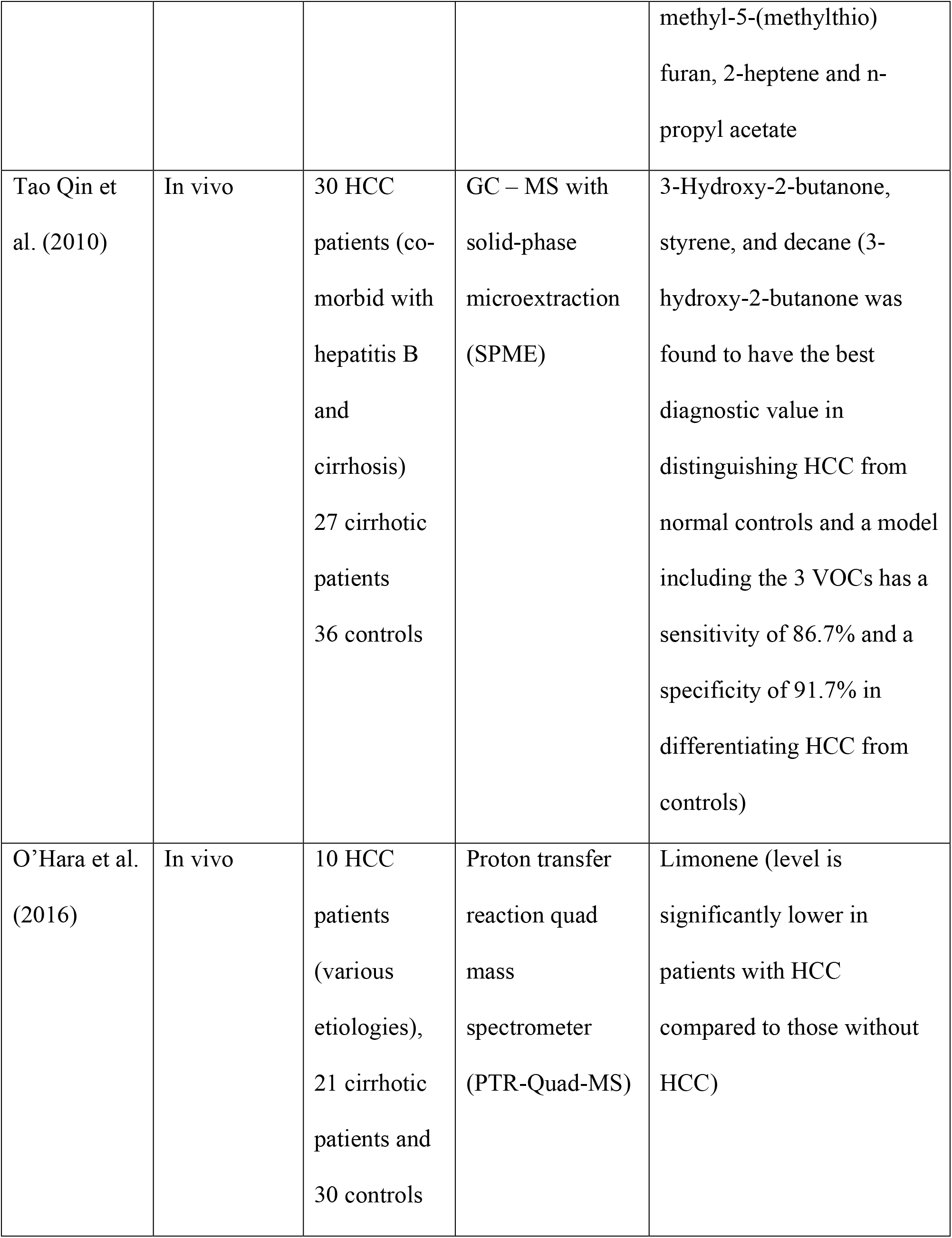
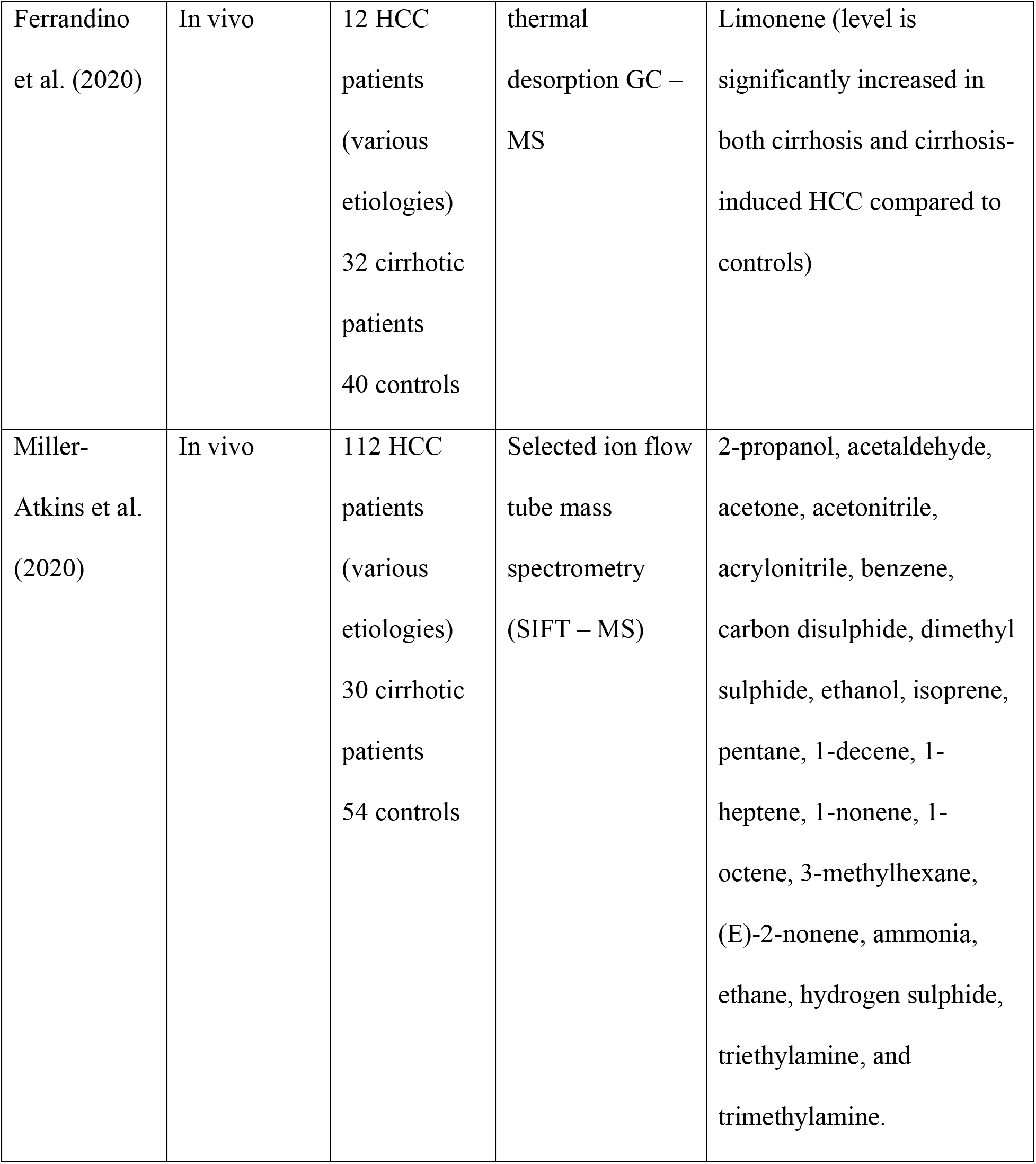
Summary of included studies

**Figure 1.**
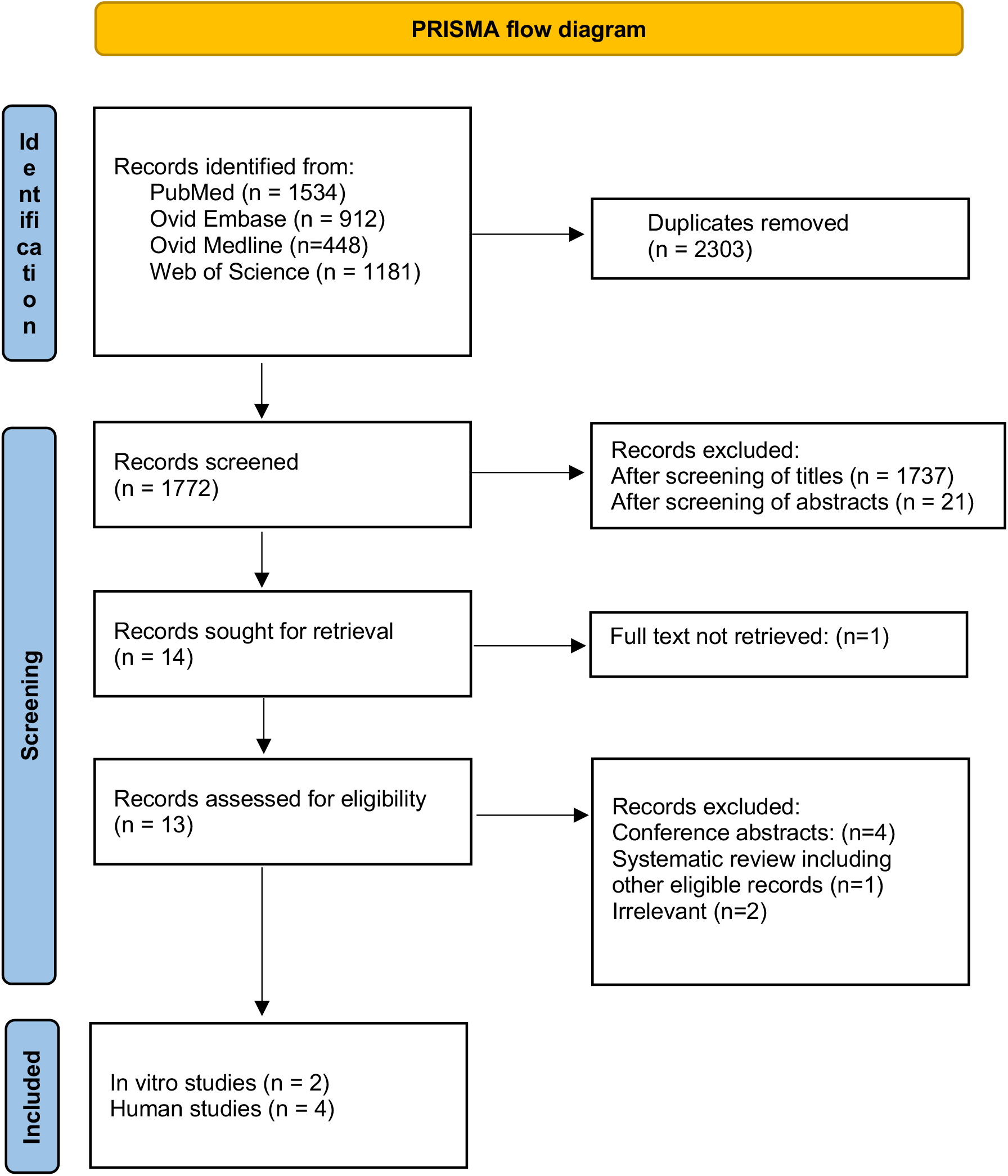
PRISMA flow diagram **Preferred Reporting items for systematic review and Meta-Analyses flowchart**.

## Results

Overall, 6 publications are included in this review. Both Amal et al. (2012) (12) and Mochalski et al. (2013) (13) generally utilized GC-MS to explore the VOCs released by in-vitro hepatocellular carcinoma cell cultures. Amal and colleagues focused on exploring characteristic VOCs changes that reflect the metastatic potential of hepatocellular carcinoma cell lines obtained by in-vivo clonal selection (12). The group utilized both GC-MS and an array of nanomaterial sensors to compare VOCs profiles of 4 low metastatic potential hepatocellular carcinoma cell lines, a high metastatic potential cell line, and a normal human immortalized hepatocyte cell line (12). They identified nine VOCs that can differentiate groups and found methane-sulfonyl chloride elevated in HCC compared to normal cell cultures, whereas 2,3 di-hydro-benzofuran is elevated in high metastatic potential HCC compared to low metastatic potential HCC. On the other hand, Mochalski et al. (2013) (13) applied GC-MS and head-space needle trap device extraction (HS-NTD) to profile the VOCs metabolized by the HepG2 liver cell line. They identified 12 metabolites released by HepG2 cells, namely 2-pentanone, 3-heptanone, 2-heptanone, 3-octane, 2-nonanone, dimethyl sulfide, ethyl methyl sulfide, 3-methyl thiophene, 2-methyl-1-(methylthio)-propane, 2-methyl-5-(methylthio) furan, 2-heptene and n-propyl acetate (13).

On the other hand, the clinical studies generally included diverse patients with comparable demographic characteristics. Qin et al. (2010) compared the VOCs released from HCC patients comorbid with hepatitis B and cirrhosis (n=30) to controls of patients with cirrhosis (n=27) and healthy volunteers (n=36) (14). The group utilized GC-MS with solid-phase microextraction (SPME) to analyse breath samples and identified 3 VOCs (3-hydroxy-2-butanone, styrene and decane) and highlighted significantly different levels of 3-hydroxy-2-butanone between the study groups (14). Qin and colleagues built a model based on the 3 identified VOCs, which showed a sensitivity of 86.7% and a specificity of 91.7% in distinguishing HCC from healthy controls (14). When applying the model to breath samples of cirrhosis patients, 33.3% were falsely diagnosed with cancer (14). On the other hand, O’Hara et al. (2016) investigated the role of breath VOCs limonene, methanol and 2-pentanone in distinguishing between liver cirrhosis (n=21), HCC (n=10) and controls (n=30) (15) They utilized proton transfer reaction quad mass spectrometer (PTR-Quad-MS) and identified significantly lower levels of limonene in patients with HCC compared to those without HCC (15). Conversely, Ferrandino et al. (2020) 16 applied thermal desorption GC-MS to measure VOCs in exhaled breath samples of 40 controls, 32 cirrhotic patients, and 12 HCC patients and found significantly increased breath limonene in both cirrhosis and cirrhosis-induced HCC compared to controls (16). Finally, Miller-Atkins et al. (2020) utilized selective ion flow tube mass spectrometry (SIFT-MS) to measure 22 VOCs in breath samples from patients with HCC (n = 112), colorectal cancer liver metastases (n = 51), cirrhosis (n = 30) and controls comprising pulmonary hypertension patients (n = 49) and subjects with no liver disease (n = 54) (17). The group highlighted the VOCs acetone, acetaldehyde and dimethyl sulfide best in differentiating cirrhosis and HCC patients, whereas (E)-2-nonene, ethane and benzene were best distinguished HCC and controls (17). Furthermore, Miller-Atkins and colleagues built a random-forest machine-learning predictive model based on VOCs and demographic variables with 85% classification accuracy. They demonstrated improved sensitivity in detecting HCC compared to AFP (73% versus 53%, respectively) (17).

Tables 1 and 2 summarise all VOCs found in the included studies. Overall, 21 VOCs have been identified by the 6 studies (Table 2). When performing pathway enrichment analysis, only 3 VOCs, namely limonene, ethanol and acetaldehyde, were present in the Kyoto Encyclopedia of Genes and Genomes (KEGG) (Fig 2). The enzyme limonene 6 monooxygenase metabolizes limonene under the control of hepatic CYP2C9 and CYP2C19. The acetaldehyde and ethanol participate in glycolysis and gluconeogenesis (Fig 2).

**Table 2.**
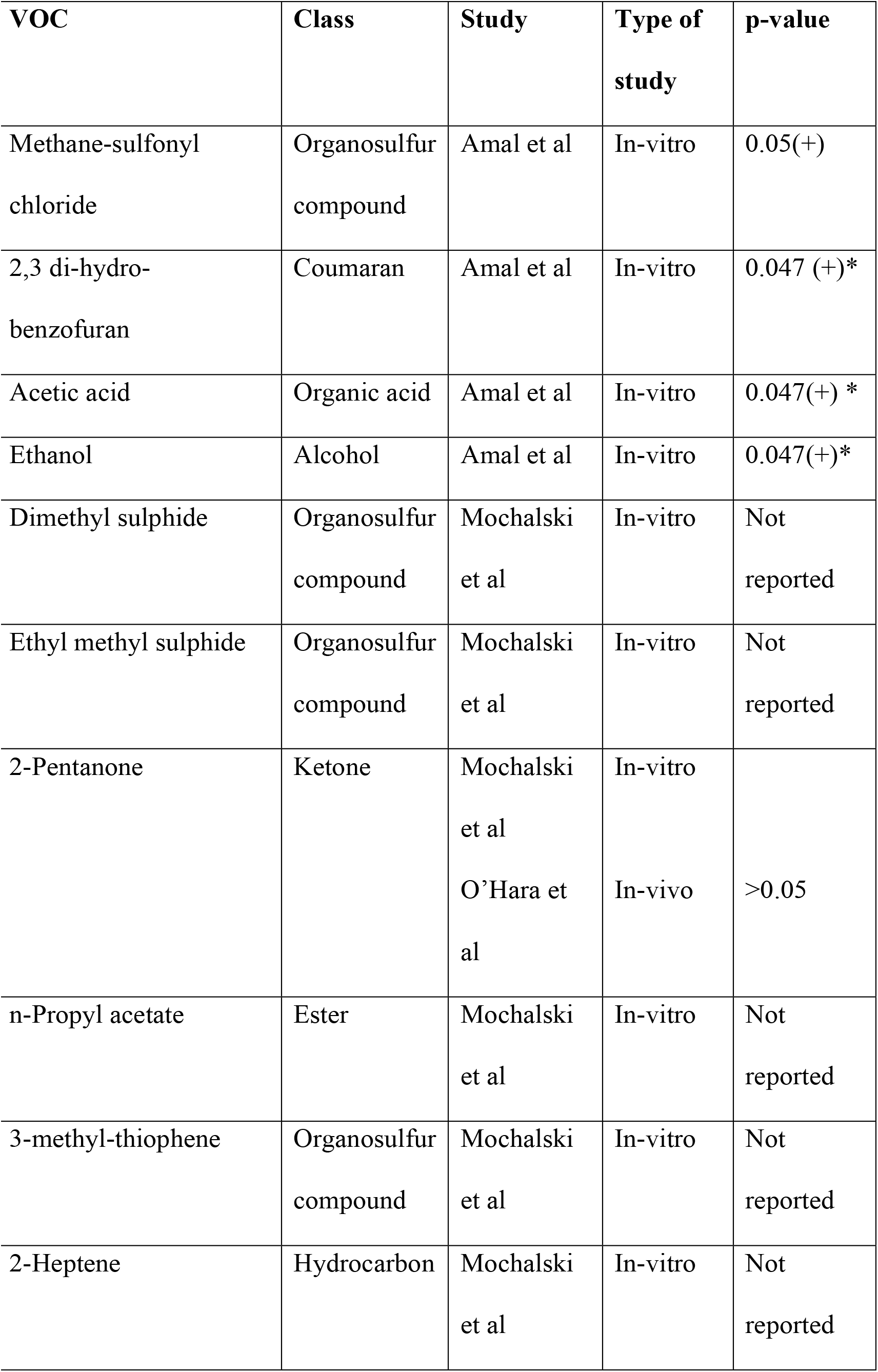
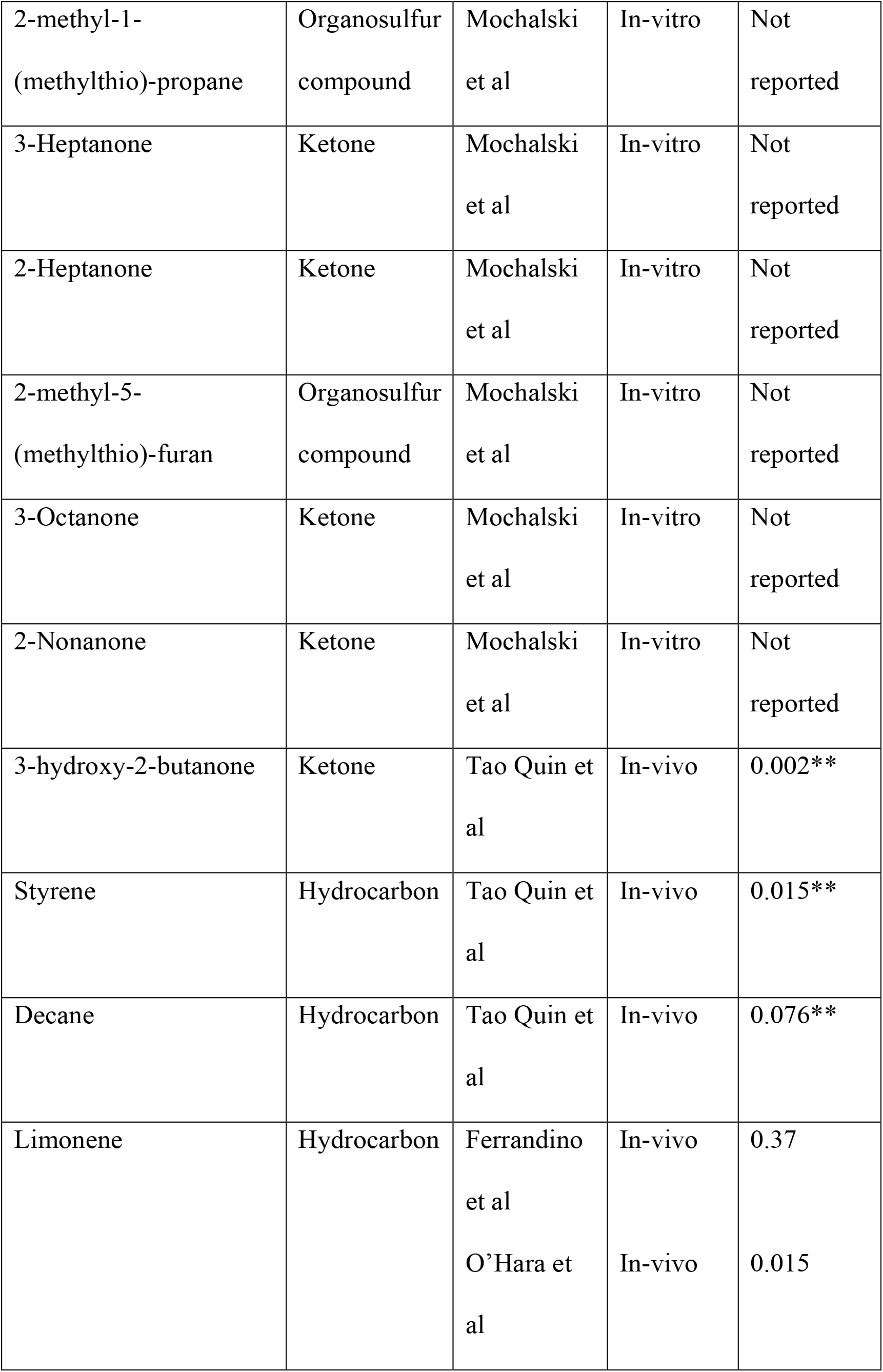
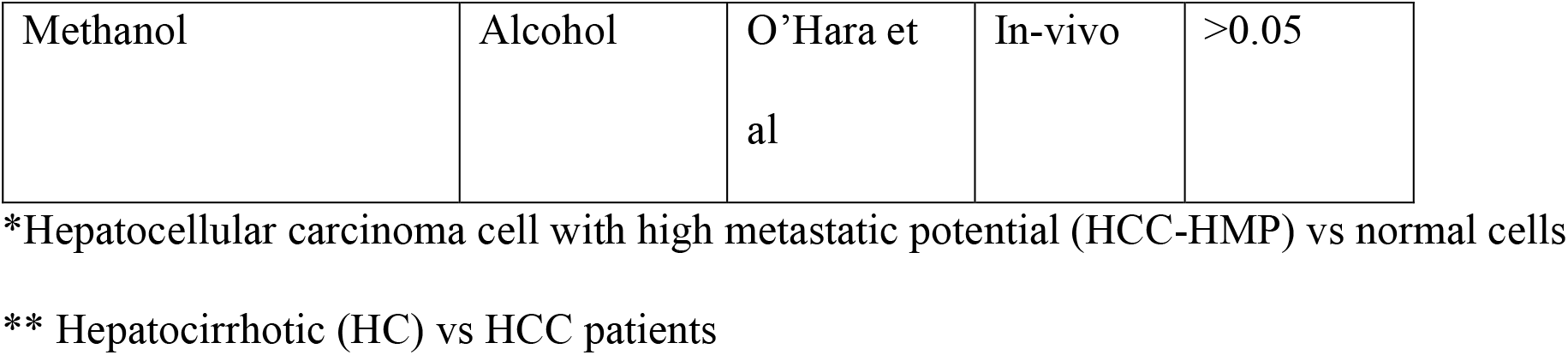
Identified volatile organic compounds (VOCs)

**Figure 2.**
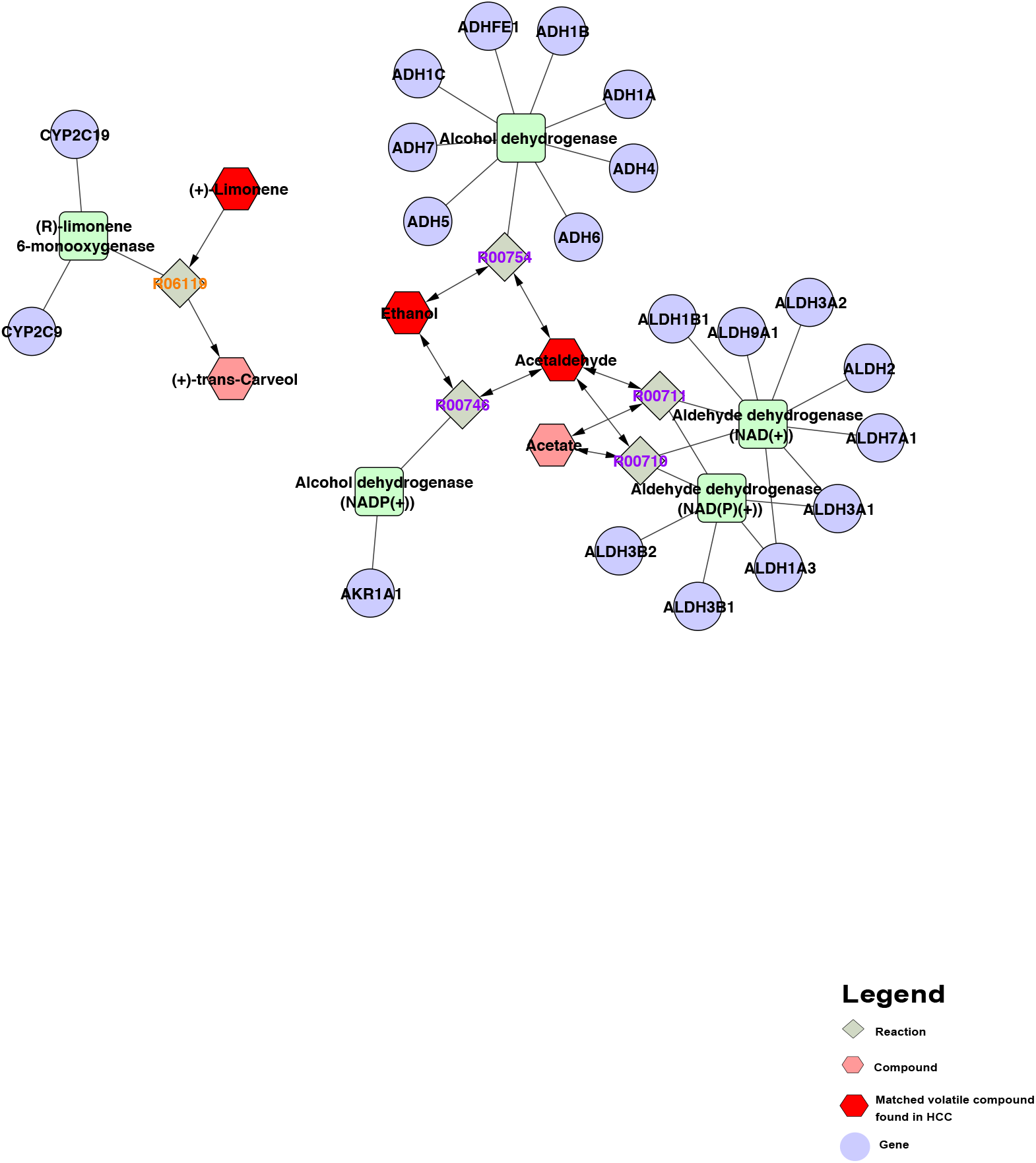
KEGG map with VOCs related to HCC annotations

## Discussion

VOCs mixtures emanated from various cells differ due to differences in cell membrane structures (12). The fundamental premise in VOCs research is that tumorigenesis causes gene and protein changes that lead to oxidative stress injury and peroxidation of the cell membrane lipids with a subsequent change in the resulting VOCs (12).

Overall, the published in-vitro VOCs research in HCC is highly variable with different cell lines and analytical methods, which are counterproductive for comparing results and generalization. Amal et al. and Mochalski et al. analysed headspace gases emitted by cultured HCC cell lines, but the HCC cell lines are dissimilar (Table 1). Generally, in-vitro experiments exclude confounding factors associated with a patient’s characteristics, such as co-morbidities, medications, smoking etc. Therefore the presence of detected substances is contributed purely to unperturbed metabolic pathways. However, the metabolic activity of isolated cell lines in tissue cultures is an oversimplification that by no means fully represents the entire dynamism of in-vivo metabolism, given the apparent lack of modulatory factors, inter-systems feedback loops and compensatory mechanisms. Therefore, the results of in-vitro research do not necessarily reflect the actual findings in in-vivo studies.

The main drawback of the in-vivo clinical HCC VOCs research is the lack of standardization. Both O’ Hara et al. (2016) (15) and Ferrandino et al. (2020) (16) investigated the role of breath limonene as a potential marker for HCC and reported contradictory results (Table 1). Limonene is an exometabolite of dietary origin that cannot be synthesized in the body and naturally occurs in citrus fruits and many vegetables (15). Furthermore, it is widely used in industry to impart citrus flavour and is an important ingredient in several perfumes and air fresheners (15). Ingested limonene is rapidly cleared in normal subjects (15), however, it tends to accumulate to a variable extent in adipose tissue in patients with liver disease owing to its highly lipophilic nature and the downregulation of the hepatic metabolizing enzymes CYP2C9 and CYP2C19 (Fig 2) (18). Due to such variability in the level of accumulated limonene, it can remain high for a few days after liver transplantation in some patients while continuing to be high for months in others (18).

Another important factor adding to the variability of the VOCs research is the analytical variability-different technologies utilized to analyse VOCs profiles in HCC research. GC-MS is the gold standard for VOCs determination and the method of choice for qualitative analysis of complex mixtures of gases (24). However, it requires laborious sample preparation and relatively lengthily analysis time therefore not capable of doing real-time analysis, is unable to perform direct quantitative analysis and requires costly standard solutions for quantitative determinations (24). GC-MS was utilised in the in-vitro studies and two in-vivo studies, whereas in the other two in-vivo studies, selected-ion flow-tube mass spectrometry (SIFT-MS) and proton transfer reaction quadrupole mass spectrometry (PTR-Quad-MS) were utilized. Combining thermal desorption or solid phase microextraction (SPME) with GC-MS reduces the ability to analyse complex gas mixtures (25). These techniques have selective affinity for certain VOCs; as such, only a fraction of the sample is analysed, which is especially counterproductive in untargeted profiling research (24).

On the other hand, direct injection mass spectrometry techniques, such as selected-ion flow-tube mass spectrometry (SIFT-MS) and proton transfer reaction mass spectrometry (PTR-MS), allow performing mass-spectrometry quantitative analysis of complex gas mixtures without the need for prior chromatographic separation (24). Therefore, these methods are capable of rapid quantitative analysis without needing pricy standards (24). There is a growing interest in PTR-MS given its high sensitivity, low detection limit, rapid real-time analysis and no requirement for a carrier gas (24). On the other hand, owing to the lack of separation in PTR-MS, it is impossible to differentiate isomers (24). Additionally, PTR-MS can only detect VOCs with proton affinity (PA) in greater water. Therefore short-chain alkanes cannot be detected (24).

On the other hand, metabolomics can help unravel the potential underlying pathophysiology of HCC. Statistically significant acetaldehyde level difference was noted, among others, by Miller-Atkins et al. (17) when comparing HCC patients to individuals with cirrhosis (Fig 2). Endogenous acetaldehyde causes alterations to the DNA strand’s structure and function, leading to carcinogenic potential (26). In healthy cells, acetaldehyde is metabolised to acetate by acetaldehyde dehydrogenase (ALDH) (27). However, it has been demonstrated that the risk of oncogenic transformation rises when ALDH capacity is diminished, particularly ALDH2, and the risk of oncogenic transformation rises (28).

Furthermore, other VOC compounds highlighted by this review were linked to cancer, such as; 3-hydroxy-2-butanone in lung cancer (29). Endogenous styrene production is also found to relate to cell toxicity (30). These findings reinforce the conceptual validity of the utility of VOCs as a diagnostic tool. However, further work is required to profile consistent biomarkers for HCC.

## Conclusions and future directions

This systematic review demonstrated a lack of unified methodology in included studies. Although no single volatile organic compound was found to be related to HCC, some light was shed on glucose metabolic pathways as a potential source of cancerous VOCs. Large population studies are required to explore the potential VOCs related to HCC.

## Data Availability

All data produced in the present work are contained in the manuscript

## Acknowledgements

The authors would like to acknowledge the supportive role of the Royal College of Surgeons of Edinburgh Liberians in conducting and verifying the search strategy

## References

1. Siegel RL, Miller KD, Jemal A. Cancer statistics, 2019. CA Cancer J Clin. 2019 Jan;69(1):7–34.

2. Sung H, Ferlay J, Siegel RL, Laversanne M, Soerjomataram I, Jemal A, et al. Global Cancer Statistics 2020: GLOBOCAN Estimates of Incidence and Mortality Worldwide for 36 Cancers in 185 Countries. CA Cancer J Clin. 2021 May;71(3):209–49.

3. Rawla P, Sunkara T, Muralidharan P, Raj JP. Update in global trends and aetiology of hepatocellular carcinoma. Contemp Oncol. 2018;22(3):141.

4. El-Serag HB. Hepatocellular carcinoma. Vol. 365, New England Journal of Medicine. Massachusetts Medical Society; 2011. p. 1118–27.

5. Llovet JM, Kelley RK, Villanueva A, Singal AG, Pikarsky E, Roayaie S, et al. Hepatocellular carcinoma. Nat Rev Dis Prim 2021 71. 2021 Jan;7(1):1–28.

6. Ioannou GN, Splan MF, Weiss NS, McDonald GB, Beretta L, Lee SP. Incidence and Predictors of Hepatocellular Carcinoma in Patients With Cirrhosis. Clin Gastroenterol Hepatol. 2007;5(8).

7. Marrero JA, Kulik LM, Sirlin CB, Zhu AX, Finn RS, Abecassis MM, et al. Diagnosis, Staging, and Management of Hepatocellular Carcinoma: 2018 Practice Guidance by the American Association for the Study of Liver Diseases. Hepatology [Internet]. 2018;68(2):723–50. Available from: http://doi.wiley.com/10.1002/hep.29913

8. Galle PR, Forner A, Llovet JM, Mazzaferro V, Piscaglia F, Raoul JL, et al. EASL Clinical Practice Guidelines: Management of hepatocellular carcinoma. J Hepatol. 2018 Jul;69(1):182–236.

9. National Guideline Centre (UK) (NICE). Cirrhosis in Over 16s: Assessment and Management. Guidelines-Editors, editor. London, United Kingdom; 2016.

10. Tzartzeva K, Obi J, Rich NE, Parikh ND, Marrero JA, Yopp A, et al. Surveillance Imaging and Alpha Fetoprotein for Early Detection of Hepatocellular Carcinoma in Patients With Cirrhosis: A Meta-analysis. Gastroenterology. 2018 May;154(6):1706-1718.e1.

11. Chan SL, Mo F, Johnson PJ, Siu DYW, Chan MHM, Lau WY, et al. Performance of serum α-fetoprotein levels in the diagnosis of hepatocellular carcinoma in patients with a hepatic mass. HPB (Oxford). 2014;16(4):366.

12. Haick H, Amal, Ding, Liu, Tisch Xu, et al. The scent fingerprint of hepatocarcinoma: in-vitro metastasis prediction with volatile organic compounds (VOCs). Int J Nanomedicine [Internet]. 2012 Jul;4135. Available from: http://www.dovepress.com/the-scent-fingerprint-of-hepatocarcinoma-in-vitro-metastasis-predictio-peer-reviewed-article-IJN

13. Mochalski P, Sponring A, King J, Unterkofler K, Troppmair J, Amann A. Release and uptake of volatile organic compounds by human hepatocellular carcinoma cells (HepG2) in vitro. Cancer Cell Int [Internet]. 2013;13(1):1. Available from: Cancer Cell International

14. Qin T, Liu H, Song Q, Song G, Wang HZ, Pan YY, et al. The screening of volatile markers for hepatocellular carcinoma. Cancer Epidemiol Biomarkers Prev. 2010;19(9):2247–53.

15. O’Hara ME, Fernández Del Río R, Holt A, Pemberton P, Shah T, Whitehouse T, et al. Limonene in exhaled breath is elevated in hepatic encephalopathy. J Breath Res. 2016;10(4).

16. Ferrandino G, Orf I, Smith R, Calcagno M, Thind AK, Debiram-Beecham I, et al. Breath Biopsy Assessment of Liver Disease Using an Exogenous Volatile Organic Compound— Toward Improved Detection of Liver Impairment. Clin Transl Gastroenterol. 2020 Sep;11(9):e00239.

17. Miller-Atkins G, Acevedo-Moreno LA, Grove D, Dweik RA, Tonelli AR, Brown JM, et al. Breath Metabolomics Provides an Accurate and Noninvasive Approach for Screening Cirrhosis, Primary, and Secondary Liver Tumors. Hepatol Commun. 2020 Jul;4(7):1041–55.

18. Fernández del Río R, O’Hara ME, Holt A, Pemberton P, Shah T, Whitehouse T, et al. Volatile Biomarkers in Breath Associated With Liver Cirrhosis - Comparisons of Pre- and Post-liver Transplant Breath Samples. EBioMedicine [Internet]. 2015;2(9):1243–50. Available from: http://dx.doi.org/10.1016/j.ebiom.2015.07.027

19. Tangerman A, Meuwese-Arends MT, Jansen JBMJ. Cause and composition of foetor hepaticus. Lancet. 1994 Feb;343(8895):483.

20. Li W, Dai W, Liu M, Long Y, Wang C, Xie S, et al. VOC biomarkers identification and predictive model construction for lung cancer based on exhaled breath analysis: research protocol for an exploratory study. BMJ Open. 2019 Aug;9(8):e028448.

21. Hua Q, Zhu Y, Liu H. Detection of volatile organic compounds in exhaled breath to screen lung cancer: a systematic review. https://doi.org/102217/fon-2017-0676. 2018 mJun;14(16):1647–62.

22. Oakley-Girvan I, Davis SW. Breath based volatile organic compounds in the detection of breast, lung, and colorectal cancers: A systematic review. Cancer Biomarkers. 2018 Jan;21(1):29–39.

23. Stravitz RT, Transplant H-L, Todd Stravitz R, Ilan Y. Potential use of metabolic breath tests to assess liver disease and prognosis: has the time arrived for routine use in the clinic? Liver Int. 2017 Mar;37(3):328–36.

24. Majchrzak T, Wojnowski W, Lubinska-Szczygeł M, Różańska A, Namieśnik J, Dymerski T. PTR-MS and GC-MS as complementary techniques for analysis of volatiles: A tutorial review. Anal Chim Acta. 2018 Dec;1035:1–13.

25. Górecki T, Yu X, Pawliszyn J. Theory of analyte extraction by selected porous polymer SPME fibres †.

26. Antonowicz S, Bodai Z, Wiggins T, Markar SR, Boshier PR, Goh YM, et al. Endogenous aldehyde accumulation generates genotoxicity and exhaled biomarkers in esophageal adenocarcinoma. Nat Commun [Internet]. 2021 Dec 5;12(1):1454. Available from: http://www.nature.com/articles/s41467-021-21800-5

27. Zakhari S. Overview: how is alcohol metabolized by the body? Alcohol Res Health [Internet]. 2006;29(4):245–54. Available from: http://www.ncbi.nlm.nih.gov/pubmed/17718403

28. Seo W, Gao Y, He Y, Sun J, Xu H, Feng D, et al. ALDH2 deficiency promotes alcohol-associated liver cancer by activating oncogenic pathways via oxidized DNA enriched extracellular vesicles. J Hepatol. 2019 Nov;71(5):1000.

29. Fu X-A, Li M, Knipp RJ, Nantz MH, Bousamra M. Noninvasive detection of lung cancer using exhaled breath. Cancer Med [Internet]. 2014 Feb;3(1):174–81. Available from: https://onlinelibrary.wiley.com/doi/10.1002/cam4.162

30. Liu C, Men X, Chen H, Li M, Ding Z, Chen G, et al. A systematic optimization of styrene biosynthesis in Escherichia coli BL21(DE3). Biotechnol Biofuels [Internet]. 2018 Dec 25;11(1):14. Available from: https://biotechnologyforbiofuels.biomedcentral.com/articles/10.1186/s13068-018-1017-z

